# Effectiveness of Human Cellular Umbilical Tissue Allograft Compared to Acellular Matrix Products in Managing Diabetic Lower Extremity Ulcers in Hospitalized Patients

**DOI:** 10.64898/2026.09.16.26363265

**Authors:** Jaideep Banerjee, Chien-Cheng Chen, Tim Styche

## Abstract

**Background:** CAMPs (cellular and acellular matrix products) can facilitate wound closure and improve clinical outcomes in patients with diabetic lower extremity ulcers (DLEU), however, there is a lack of comparative data between different CAMPs. The objective of this study was to use real-world evidence to compare the clinical and economic outcomes of hospital care for DLEU patients treated with a minimally manipulated human umbilical tissue (STRAVIX^◊^) or other acellular CAMPs.

**Methods:** This retrospective analysis was conducted on patients with DLEU treated with the CAMPs using data from the Premier PINC AI Healthcare Database. Each patient’s earliest inpatient encounter was designated as the index admission. The analysis groups included a cellular human umbilical tissue allograft (STRAVIX) and four different acellular CAMPs (aCAMPs). To balance baseline covariates, inverse probability of treatment weighting (IPTW) was applied. Subsequently, IPTW-adjusted generalized linear models were utilized to estimate healthcare-related outcomes and hospital-related costs.

**Results:** 2,626 DLEUs patients with STRAVIX and 10,002 DLEUs patients with aCAMPs were identified. Compared to the patients treated with aCAMPs, the patients who had STRAVIX in addition to standard of care had significantly lower odds of having an infection reaction, dehiscence, minor and major amputations, after index admission discharge. The number of DLEU related follow-up visits was 17-23% lower, debridement related visits were 19-23% lower and the incidence of all-cause readmission was 22-25% lower in Stravix patients compared to aCAMP patients. The length of hospital stay days was 3% lower and the cost of all-cause visits was 20-25% lower in STRAVIX treated patients compared to aCAMP patients.

**Conclusion:** This large data set demonstrates improved clinical outcomes in patients who had STRAVIX in addition to standard surgical management, when compared to certain acellular CAMPs.

## Introduction

Diabetic lower extremity ulcers (DLEUs) continue to pose a serious medical and financial burden to society. In the United States, approximately 40.1 million people are living with diabetes amongst whom 19-34% has a risk of developing an ulcer[1, 2]. Lower extremity amputation is the most feared complication in people with DLEUs (100000 amputations annually in the US) and produces a significant physical, psychosocial and economic burden[3, 4]. Of patients having a DLEU, about 21% may have an amputation and the 5-year mortality rates can range from 54% for a minor amputation and may exceed 90% after a major amputation[1, 5].

Treatment depends on the severity of the wound which starts with standard of care including debridement, infection control, moisture balance[6]. With standard-of-care wound management, pooled Level 1 evidence shows that only about 33% of DLEUs achieve complete healing within 12 to 24 weeks, leaving majority at risk for ongoing complications[6, 7]. These hard-to-heal patients therefore may benefit from advanced support such as skin substitutes or CAMPs. Over the years, CAMPs have been proven to improve limb salvage and significantly improve patient outcomes by decreasing the incidence of major amputations, reducing emergency department visits and facilitating wound closure by improving healing rates and reducing infection risks, ultimately lowering healthcare costs[8, 9].

Amniotic membranes originating from the human placenta or the umbilical cord have emerged as a promising adjunct to standard of care and facilitate wound care. This specialized tissue is ethically sourced from consenting individuals during planned cesarean sections, after which it undergoes careful processing designed to preserve its native components. The primary component of the tissue allograft is extracellular matrix and depending on processing techniques, retains growth factors and native cells of the tissue. A recently published, large-scale, propensity score-matched analysis presented evidence of amniotic tissue allografts being a viable alternative to conventional bioengineered skin substitutes. The analysis demonstrated significantly reduced rates of infections, dehiscence, and a decreased need for subsequent autografting, suggesting that the properties of these tissues may be especially beneficial in more complex wound environments[10]. An observational hierarchical cluster analysis of risk ratios and time to closure also trended towards a better clinical outcome with amniotic allografts[11].

One such commercially available product in the USA is a strong and durable allograft, processed from the human umbilical cord. The allograft is available either as cryopreserved or lyopreserved (shelf stable), together classified here as the STRAVIX^◊^ family. STRAVIX is minimally manipulated and retains the hyaluronic acid rich extracellular matrix, native growth factors and cells of the tissue and can be used as a wrap, cover or barrier on wounds of different etiologies including DLEUs. Cryopreserved and lyopreserved amnion have been reported to be clinically equivalent[12]. These allografts are thick, suture-able and can be effective in covering large complex wounds. In a prospective randomized clinical trial, in challenging complex wounds with exposed deep structures, high rates of granulation were observed with two applications of the allograft. In a prospective pilot study in complex acute and chronic wounds, 80% of evaluable wounds achieved 100% granulation with a median time to complete granulation of 13 days[13]. In a case series evaluated in lower-extremity gas gangrene, authors reported shorter hospitalization and fewer complications with all wounds closed in approximately 3.3 months[14]. However, comparative data for STRAVIX against other CAMPs is not available. This retrospective registry analysis for the first time reports clinical outcomes compared to some of the commonly available CAMPs in the inpatient DLEU population.

## Methods

### Data Source

This retrospective cohort study used the Premier PINC AI Healthcare Database (PHD; Premier Inc.) from 1 January 2017 to 30 September 2025. The PHD is a large, U.S. hospital-based, service-level, all-payer database that includes information on inpatient discharges from geographically diverse nonprofit, nongovernmental, community, and teaching hospitals and health systems in rural and urban settings. The PHD contains more than 135 million visits, with over 13 million visits per year since 2012, representing approximately 25% of annual U.S. inpatient admissions[15]. The PHD has been certified as de-identified, and all data complies with the Health Insurance Portability and Accountability Act[15].

### Study Population

Patients with diabetic lower extremity ulcers (DLEU) who received STRAVIX (manufactured by Smith & Nephew) or other acellular CAMP products (aCAMPs), were identified from the PHD. Eligible patients were required to have at least one inpatient encounter with a DLEU diagnosis, defined using ICD-10-CM diagnosis codes for diabetes mellitus (E08X–E13X) and lower extremity ulcer (L97X), during the identification period from 1 January 2017 to 30 September 2025.

Treatment exposure was identified using hospital billing records, charge-description keyword searches, and Current Procedural Terminology (CPT)/Healthcare Common Procedure Coding System (HCPCS) Codes. STRAVIX encounters were identified by billing records containing the keyword “STRAVIX,” excluding records containing “GRAFIX,” or by HCPCS codes Q4132 or Q4133. aCAMP encounters were identified using product-specific billing descriptions or HCPCS codes, including Q4118, Q4119, Q4166, Q4158, A2019, Q4104, Q4105, Q4108, Q4114, Q4172, Q4195, Q4196, and Q4187.

For each patient, the earliest qualifying inpatient encounter was designated as the index admission, and the admission date of that encounter was defined as the index date. Patients were included if they were aged ≥18 years and had at least 6 months of observable baseline data before the index admission and at least 6 months of follow-up after discharge. Patients were excluded if they received the same treatment during the baseline period, received both STRAVIX and aCAMP products during the baseline or follow-up period, or had missing or unknown gender or race information. **(Figure 1)**

**Figure 1.**
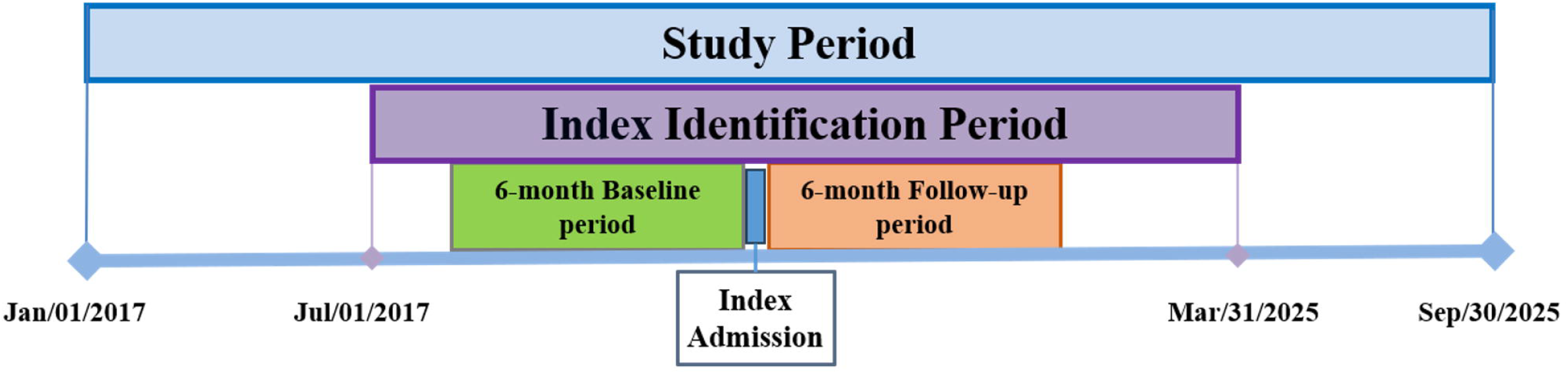
Study design

### Demographics and Baseline Characteristics

Baseline characteristics included patient demographics, clinical characteristics, and hospital-related variables. Patient characteristics included age, gender, race, year of index admission, ulcer location, use of negative pressure wound therapy (NPWT), use of hyperbaric oxygen therapy (HBOT), and comorbidities, including hypertension, obesity, tobacco use history, diabetes mellitus, myocardial infarction, congestive heart failure, depression, sepsis, cellulitis, osteomyelitis, and Charlson Comorbidity Index. Comorbidities were assessed using diagnosis records from the 6-month baseline period through the index admission. Ulcer location, NPWT use, and HBOT use were assessed during the index admission using diagnosis, HCPCS, or CPT codes, as applicable. Hospital-related variables included region, bed size, rural or urban location, teaching status, and payer type.

### Study Outcomes

Study outcomes included clinical, healthcare resource utilization (HRU), and economic endpoints assessed within 1, 3, and 6 months after discharge from the index admission. Clinical outcomes included all-cause visits, DLEU-related visits, debridement-related visits (CPT: 11042-11047, 97597, 97598), infection (ICD-10-CM: T81.4X) or dehiscence-related visits (ICD-10-CM: T81.3X), and minor (ICD-10-CM: Z89.4X) or major amputations (ICD-10-CM: Z89.5X, Z89.6X). Incidence was defined as the proportion of patients with at least one qualifying event during each follow-up period. HRU outcomes included length of stay during the index admission and visit frequency during follow-up, reported as the number of visits per 100 patients. Economic outcomes included total hospital costs during the index admission and total costs of all-cause visits during follow-up. All costs were inflation-adjusted to 2025 U.S. dollars using the medical care component of the Consumer Price Index.

### Statistical Analysis

Stabilized inverse probability of treatment weighting (sIPTW) based on propensity scores was used to balance measured demographics and baseline characteristics between the STRAVIX and aCAMP cohorts while retaining all eligible patients in the analytic sample. Propensity scores estimated each patient’s probability of receiving STRAVIX conditional on measured baseline covariates. Stabilized weights were calculated as the marginal probability of the observed treatment divided by the propensity score for patients receiving STRAVIX and divided by 1 minus the propensity score for patients receiving aCAMP products. Covariate balance before and after weighting was assessed using standardized mean differences (SMDs). Adequate balance was defined as SMD less than 10. Weighted regression models were used to compare outcomes between cohorts after sIPTW adjustment. Weighted logistic regression models were used for binary incidence outcomes, including all-cause visits, DLEU-related visits, debridement-related visits, infection- or dehiscence-related visits, and amputations, with treatment effects reported as odds ratios (ORs) and 95% confidence intervals (CIs). Weighted generalized linear models with a negative binomial distribution were used for length of stay and visit frequency outcomes, with treatment effects reported as rate ratios (RRs) and 95% CIs. Weighted generalized linear models with a gamma distribution were used for index-admission costs, and models with a Tweedie distribution were used for total all-cause follow-up costs, with treatment effects reported as ratios of means and 95% CIs. All analyses were conducted using SAS for Windows version 9.4 (SAS Institute Inc.). Two-sided p-values <0.05 were considered statistically significant.

### Ethical approval and patient consent

Obtaining informed consent was not necessary as all patient records were deidentified, and all data were compliant with the Health Insurance Portability and Accountability Act[16]. Consequently, this study was exempt from Institutional Review Board approval.

## Results

### Patient selection

Patients with at least 1 inpatient encounter with a DLEU diagnosis during the identification period resulted in 603,875 subjects. Retaining patients with either a STRAVIX or aCAMP encounter and applying exclusion criteria, resulted in a final cohort of 12,628 subjects of which 2,626 subjects had a STRAVIX application while 10,002 subjects had aCAMP application **(Figure 2)**.

**Figure 2.**
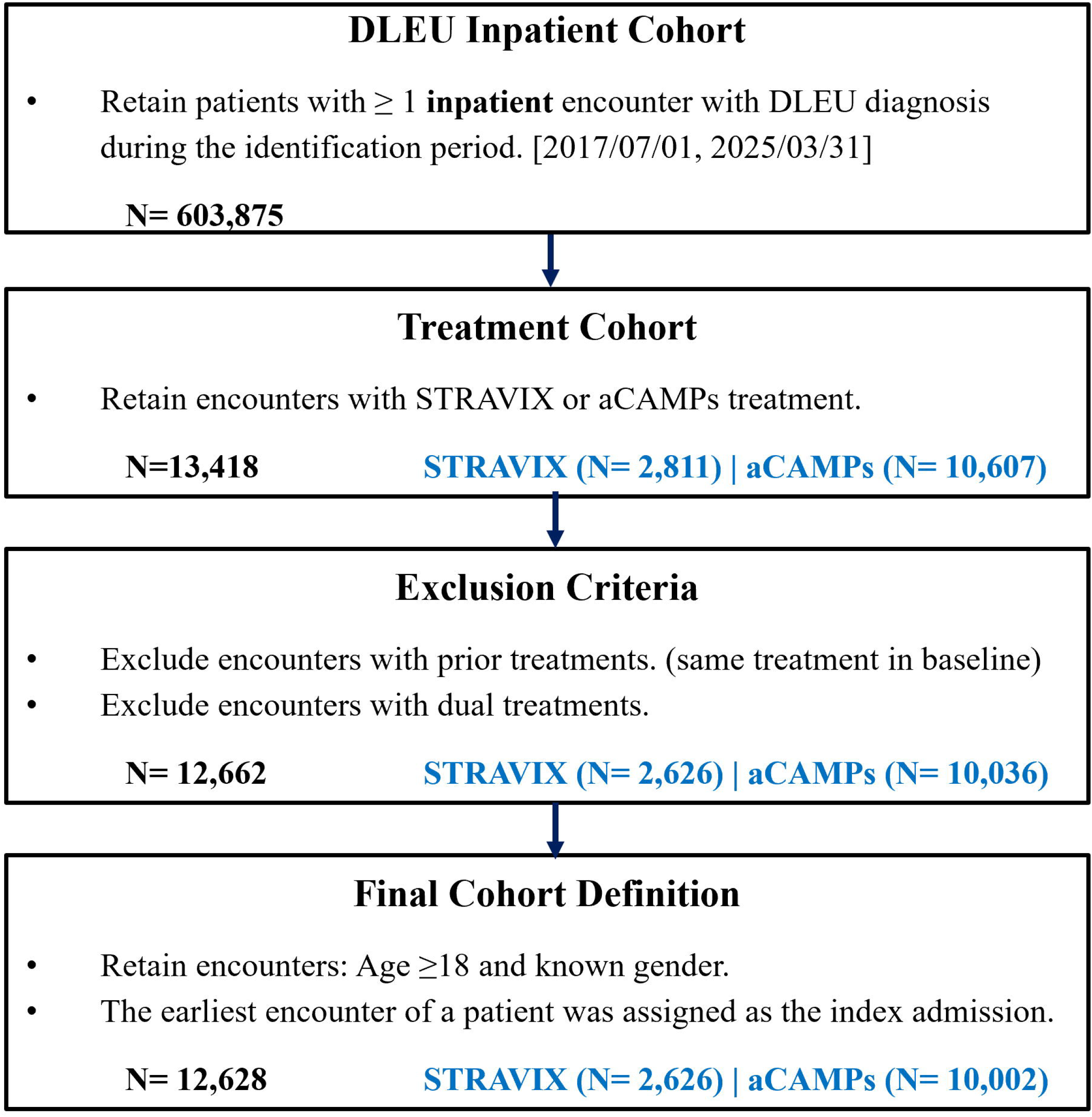
Patient Selection Flowchart

### Patient Demographics

Table 1 characterizes baseline demographics, hospital characteristics and clinical comorbidities. Before weighting, the STRAVIX and aCAMPs cohorts were generally similar with respect to demographic and clinical characteristics. However, compared with the aCAMPs cohort, STRAVIX-treated patients were more frequently treated in smaller and rural hospitals, were more commonly from the Northeast and West regions, had lower NPWT use during the index admission, and had a higher proportion of ulcers located at the ankle, heel, or foot. After application of sIPTW, all baseline characteristics were well balanced between cohorts, with SMD less than 10, while retaining all eligible patients in the analytic sample.

**Table 1:**
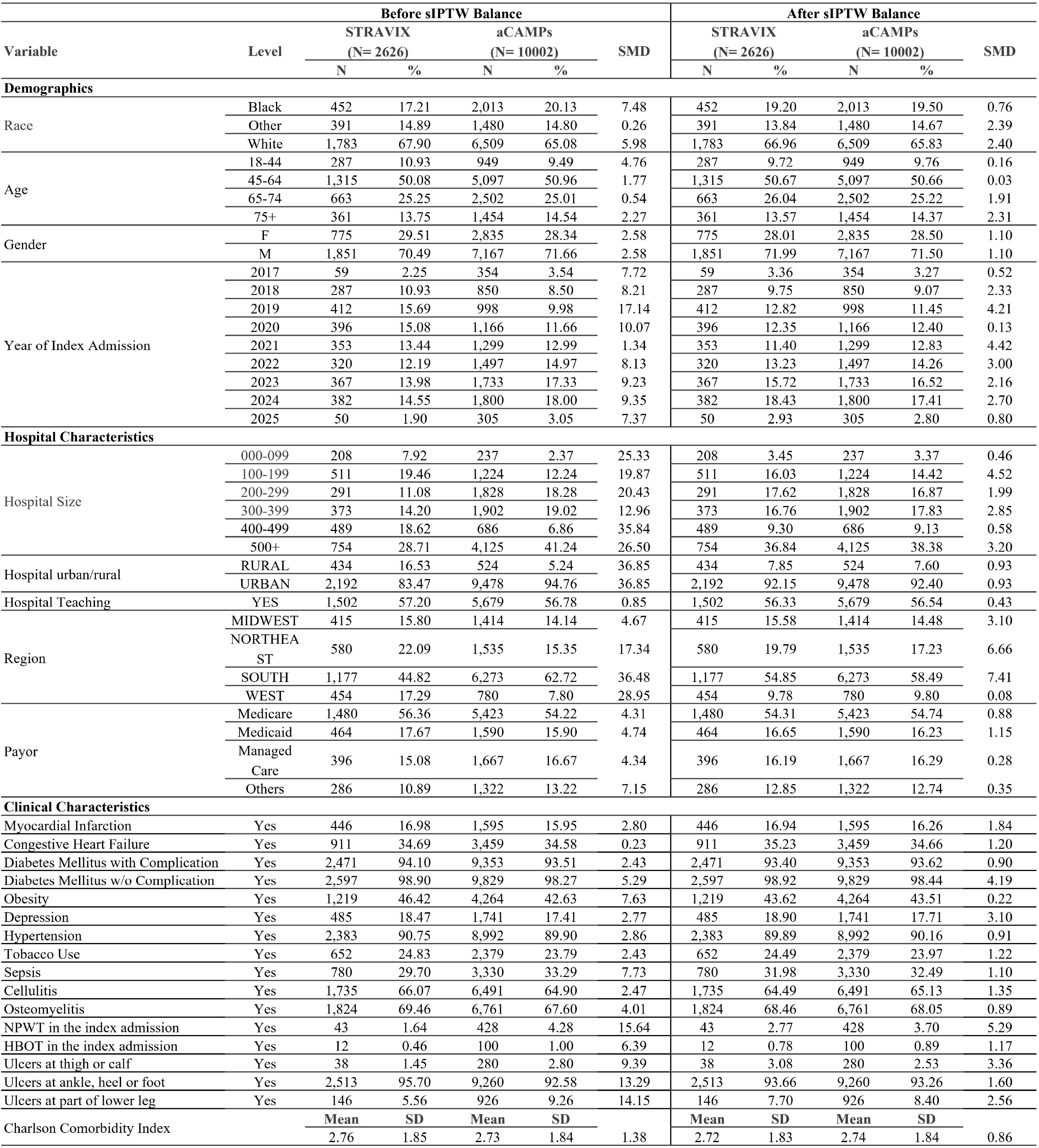
Baseline Characteristics before and after IPTW balancing.

| Before sIPTW Balance |  |  |  |  |  |  | After sIPTW Balance |  |  |  |  |
| --- | --- | --- | --- | --- | --- | --- | --- | --- | --- | --- | --- |
| Variable | Level | STRAVIX |  | aCAMPs |  | SMD | STRAVIX |  | aCAMPs |  | SMD |
|  |  | (N= 2626) |  | (N= 10002) |  |  | (N= 2626) |  | (N= 10002) |  |  |
|  |  | N | % | N | % |  | N | % | N | % |  |
| Demographics |  |  |  |  |  |  |  |  |  |  |  |
| Race | Black | 452 | 17.21 | 2,013 | 20.13 | 7.48 | 452 | 19.20 | 2,013 | 19.50 | 0.76 |
|  | Other | 391 | 14.89 | 1,480 | 14.80 | 0.26 | 391 | 13.84 | 1,480 | 14.67 | 2.39 |
|  | White | 1,783 | 67.90 | 6,509 | 65.08 | 5.98 | 1,783 | 66.96 | 6,509 | 65.83 | 2.40 |
| Age | 18-44 | 287 | 10.93 | 949 | 9.49 | 4.76 | 287 | 9.72 | 949 | 9.76 | 0.16 |
|  | 45-64 | 1,315 | 50.08 | 5,097 | 50.96 | 1.77 | 1,315 | 50.67 | 5,097 | 50.66 | 0.03 |
|  | 65-74 | 663 | 25.25 | 2,502 | 25.01 | 0.54 | 663 | 26.04 | 2,502 | 25.22 | 1.91 |
|  | 75+ | 361 | 13.75 | 1,454 | 14.54 | 2.27 | 361 | 13.57 | 1,454 | 14.37 | 2.31 |
| Gender | F | 775 | 29.51 | 2,835 | 28.34 | 2.58 | 775 | 28.01 | 2,835 | 28.50 | 1.10 |
|  | M | 1,851 | 70.49 | 7,167 | 71.66 | 2.58 | 1,851 | 71.99 | 7,167 | 71.50 | 1.10 |
| Year of Index Admission | 2017 | 59 | 2.25 | 354 | 3.54 | 7.72 | 59 | 3.36 | 354 | 3.27 | 0.52 |
|  | 2018 | 287 | 10.93 | 850 | 8.50 | 8.21 | 287 | 9.75 | 850 | 9.07 | 2.33 |
|  | 2019 | 412 | 15.69 | 998 | 9.98 | 17.14 | 412 | 12.82 | 998 | 11.45 | 4.21 |
|  | 2020 | 396 | 15.08 | 1,166 | 11.66 | 10.07 | 396 | 12.35 | 1,166 | 12.40 | 0.13 |
|  | 2021 | 353 | 13.44 | 1,299 | 12.99 | 1.34 | 353 | 11.40 | 1,299 | 12.83 | 4.42 |
|  | 2022 | 320 | 12.19 | 1,497 | 14.97 | 8.13 | 320 | 13.23 | 1,497 | 14.26 | 3.00 |
|  | 2023 | 367 | 13.98 | 1,733 | 17.33 | 9.23 | 367 | 15.72 | 1,733 | 16.52 | 2.16 |
|  | 2024 | 382 | 14.55 | 1,800 | 18.00 | 9.35 | 382 | 18.43 | 1,800 | 17.41 | 2.70 |
|  | 2025 | 50 | 1.90 | 305 | 3.05 | 7.37 | 50 | 2.93 | 305 | 2.80 | 0.80 |
| Hospital Characteristics |  |  |  |  |  |  |  |  |  |  |  |
| Hospital Size | 000-099 | 208 | 7.92 | 237 | 2.37 | 25.33 | 208 | 3.45 | 237 | 3.37 | 0.46 |
|  | 100-199 | 511 | 19.46 | 1,224 | 12.24 | 19.87 | 511 | 16.03 | 1,224 | 14.42 | 4.52 |
|  | 200-299 | 291 | 11.08 | 1,828 | 18.28 | 20.43 | 291 | 17.62 | 1,828 | 16.87 | 1.99 |
|  | 300-399 | 373 | 14.20 | 1,902 | 19.02 | 12.96 | 373 | 16.76 | 1,902 | 17.83 | 2.85 |
|  | 400-499 | 489 | 18.62 | 686 | 6.86 | 35.84 | 489 | 9.30 | 686 | 9.13 | 0.58 |
|  | 500+ | 754 | 28.71 | 4,125 | 41.24 | 26.50 | 754 | 36.84 | 4,125 | 38.38 | 3.20 |
| Hospital urban/rural | RURAL | 434 | 16.53 | 524 | 5.24 | 36.85 | 434 | 7.85 | 524 | 7.60 | 0.93 |
|  | URBAN | 2,192 | 83.47 | 9,478 | 94.76 | 36.85 | 2,192 | 92.15 | 9,478 | 92.40 | 0.93 |
| Hospital Teaching | YES | 1,502 | 57.20 | 5,679 | 56.78 | 0.85 | 1,502 | 56.33 | 5,679 | 56.54 | 0.43 |
| Region | MIDWEST | 415 | 15.80 | 1,414 | 14.14 | 4.67 | 415 | 15.58 | 1,414 | 14.48 | 3.10 |
|  | NORTHEAST | 580 | 22.09 | 1,535 | 15.35 | 17.34 | 580 | 19.79 | 1,535 | 17.23 | 6.66 |
|  | SOUTH | 1,177 | 44.82 | 6,273 | 62.72 | 36.48 | 1,177 | 54.85 | 6,273 | 58.49 | 7.41 |
|  | WEST | 454 | 17.29 | 780 | 7.80 | 28.95 | 454 | 9.78 | 780 | 9.80 | 0.08 |
| Payor | Medicare | 1,480 | 56.36 | 5,423 | 54.22 | 4.31 | 1,480 | 54.31 | 5,423 | 54.74 | 0.88 |
|  | Medicaid | 464 | 17.67 | 1,590 | 15.90 | 4.74 | 464 | 16.65 | 1,590 | 16.23 | 1.15 |
|  | Managed Care | 396 | 15.08 | 1,667 | 16.67 | 4.34 | 396 | 16.19 | 1,667 | 16.29 | 0.28 |
|  | Others | 286 | 10.89 | 1,322 | 13.22 | 7.15 | 286 | 12.85 | 1,322 | 12.74 | 0.35 |
| Clinical Characteristics |  |  |  |  |  |  |  |  |  |  |  |
| Myocardial Infarction | Yes | 446 | 16.98 | 1,595 | 15.95 | 2.80 | 446 | 16.94 | 1,595 | 16.26 | 1.84 |
| Congestive Heart Failure | Yes | 911 | 34.69 | 3,459 | 34.58 | 0.23 | 911 | 35.23 | 3,459 | 34.66 | 1.20 |
| Diabetes Mellitus with Complication | Yes | 2,471 | 94.10 | 9,353 | 93.51 | 2.43 | 2,471 | 93.40 | 9,353 | 93.62 | 0.90 |
| Diabetes Mellitus w/o Complication | Yes | 2,597 | 98.90 | 9,829 | 98.27 | 5.29 | 2,597 | 98.92 | 9,829 | 98.44 | 4.19 |
| Obesity | Yes | 1,219 | 46.42 | 4,264 | 42.63 | 7.63 | 1,219 | 43.62 | 4,264 | 43.51 | 0.22 |
| Depression | Yes | 485 | 18.47 | 1,741 | 17.41 | 2.77 | 485 | 18.90 | 1,741 | 17.71 | 3.10 |
| Hypertension | Yes | 2,383 | 90.75 | 8,992 | 89.90 | 2.86 | 2,383 | 89.89 | 8,992 | 90.16 | 0.91 |
| Tobacco Use | Yes | 652 | 24.83 | 2,379 | 23.79 | 2.43 | 652 | 24.49 | 2,379 | 23.97 | 1.22 |
| Sepsis | Yes | 780 | 29.70 | 3,330 | 33.29 | 7.73 | 780 | 31.98 | 3,330 | 32.49 | 1.10 |
| Cellulitis | Yes | 1,735 | 66.07 | 6,491 | 64.90 | 2.47 | 1,735 | 64.49 | 6,491 | 65.13 | 1.35 |
| Osteomyelitis | Yes | 1,824 | 69.46 | 6,761 | 67.60 | 4.01 | 1,824 | 68.46 | 6,761 | 68.05 | 0.89 |
| NPWT in the index admission | Yes | 43 | 1.64 | 428 | 4.28 | 15.64 | 43 | 2.77 | 428 | 3.70 | 5.29 |
| HBOT in the index admission | Yes | 12 | 0.46 | 100 | 1.00 | 6.39 | 12 | 0.78 | 100 | 0.89 | 1.17 |
| Ulcers at thigh or calf | Yes | 38 | 1.45 | 280 | 2.80 | 9.39 | 38 | 3.08 | 280 | 2.53 | 3.36 |
| Ulcers at ankle, heel or foot | Yes | 2,513 | 95.70 | 9,260 | 92.58 | 13.29 | 2,513 | 93.66 | 9,260 | 93.26 | 1.60 |
| Ulcers at part of lower leg | Yes | 146 | 5.56 | 926 | 9.26 | 14.15 | 146 | 7.70 | 926 | 8.40 | 2.56 |
| Charlson Comorbidity Index |  | Mean | SD | Mean | SD | 1.38 | Mean | SD | Mean | SD | 0.86 |
|  |  | 2.76 | 1.85 | 2.73 | 1.84 |  | 2.72 | 1.83 | 2.74 | 1.84 |  |

### Clinical outcomes after discharge

Patients who had an application of STRAVIX had significantly lower odds of having an infection reaction compared to patients who received aCAMPs, within 1 month, 3 month and 6 months, after index admission discharge. The odds of dehiscence in patients who had an application of STRAVIX was also significantly lower at 3 and 6 months after index admission discharge compared to patients who received aCAMPs. The odds of having a minor amputation of the toe, foot, or ankle was significantly lower at 1, 3, and 6 months in patients who received STRAVIX. Major amputation of leg below knee or above knee had lower odds at 6 months in patients who received STRAVIX. Overall, any all-cause visits had significantly lower odds in patients who received STRAVIX compared to those who received aCAMPS. **(Table 2)**.

**Table 2:**
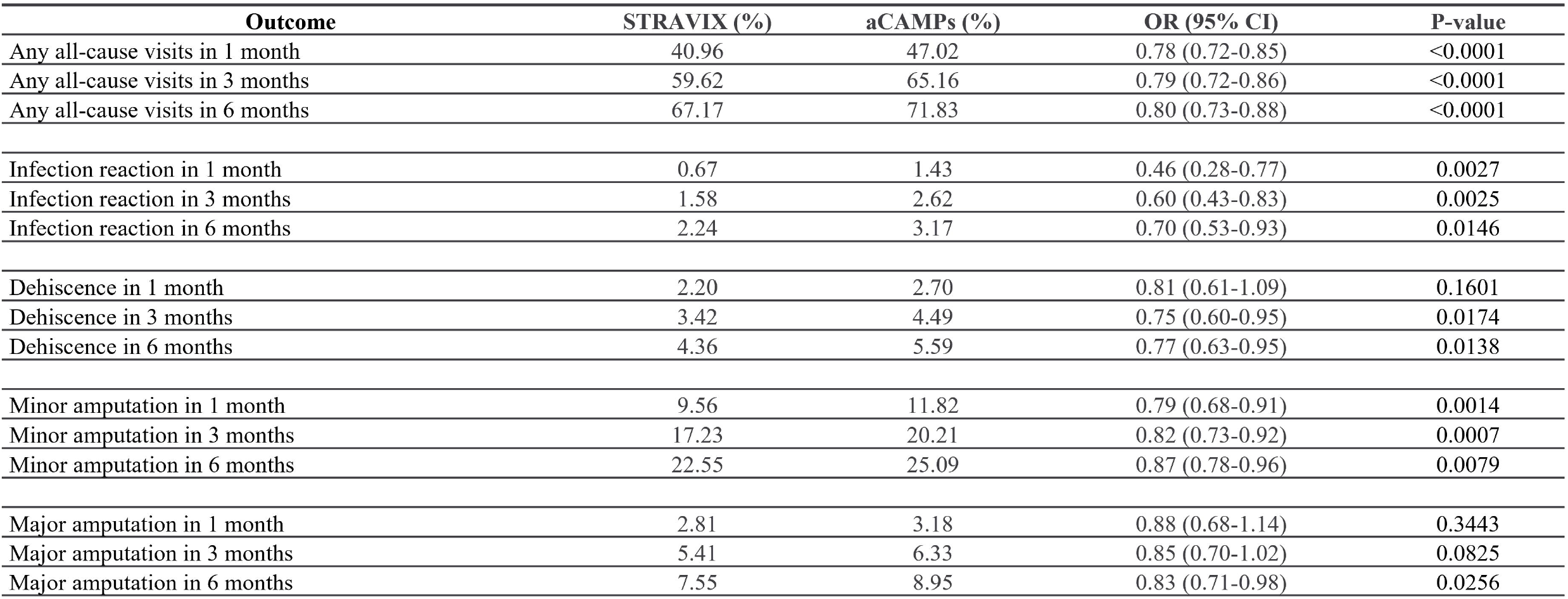
Clinical Outcomes after discharge.

| Outcome | STRAVIX (%) | aCAMPs (%) | OR (95% CI) | P-value |
| --- | --- | --- | --- | --- |
| Any all-cause visits in 1 month | 40.96 | 47.02 | 0.78 (0.72-0.85) | <0.0001 |
| Any all-cause visits in 3 months | 59.62 | 65.16 | 0.79 (0.72-0.86) | <0.0001 |
| Any all-cause visits in 6 months | 67.17 | 71.83 | 0.80 (0.73-0.88) | <0.0001 |
| Infection reaction in 1 month | 0.67 | 1.43 | 0.46 (0.28-0.77) | 0.0027 |
| Infection reaction in 3 months | 1.58 | 2.62 | 0.60 (0.43-0.83) | 0.0025 |
| Infection reaction in 6 months | 2.24 | 3.17 | 0.70 (0.53-0.93) | 0.0146 |
| Dehiscence in 1 month | 2.20 | 2.70 | 0.81 (0.61-1.09) | 0.1601 |
| Dehiscence in 3 months | 3.42 | 4.49 | 0.75 (0.60-0.95) | 0.0174 |
| Dehiscence in 6 months | 4.36 | 5.59 | 0.77 (0.63-0.95) | 0.0138 |
| Minor amputation in 1 month | 9.56 | 11.82 | 0.79 (0.68-0.91) | 0.0014 |
| Minor amputation in 3 months | 17.23 | 20.21 | 0.82 (0.73-0.92) | 0.0007 |
| Minor amputation in 6 months | 22.55 | 25.09 | 0.87 (0.78-0.96) | 0.0079 |
| Major amputation in 1 month | 2.81 | 3.18 | 0.88 (0.68-1.14) | 0.3443 |
| Major amputation in 3 months | 5.41 | 6.33 | 0.85 (0.70-1.02) | 0.0825 |
| Major amputation in 6 months | 7.55 | 8.95 | 0.83 (0.71-0.98) | 0.0256 |

### Resource utilization after discharge

Patients who received an application of STRAVIX had 23% lower DLEU related visits within 1 month (0.36 visits per STRAVIX patient, compared to 0.46 visits per aCAMP patient), 19% lower by 3 months for STRAVIX patients (1.03 visits per patient compared to 1.27 visits per aCAMP patient) and 17% lower at 6 months for STRAVIX patients (1.69 visits per patient compared to 2.02 visits per aCAMP patient). Patients who received a STRAVIX application had 19% lower debridement related visits within 1 month (0.22 visits per STRAVIX patient compared to 0.27 visits per aCAMP patient), 22% lower by 3 months for STRAVIX patients (0.60 visits per patient compared to 0.77 visits per aCAMP patient) and 23% lower at 6 months for STRAVIX patients (0.94 visits per patient compared to 1.22 visits per aCAMP patient). Patients who received an application of STRAVIX also had 25% lower rates of all-cause readmission within 1 month in STRAVIX patients (0.85 visits per patient compared to 1.14 visits per aCAMP patient), 24% lower at 3 months for STRAVIX patients (2.14 visits per patient compared to 2.81 visits per aCAMP patient) and 22% lower at 6 months for STRAVIX patients (3.45 visits per patient compared to 4.42 visits per aCAMP patient) **(Table 3)**.

**Table 3:** Resource Utilization after Discharge.

| **Outcome** | **STRAVIX**<br>**(per 100 pts)** | **aCAMPs**<br>**(per 100 pts)** | **Rate Ratio (95% CI)** | **P-value** |
| --- | --- | --- | --- | --- |
| All-cause visits in 1 month | 85.1 | 113.7 | 0.75 (0.69-0.81) | <0.0001 |
| All-cause visits in 3 months | 214.2 | 280.6 | 0.76 (0.72-0.82) | <0.0001 |
| All-cause visits in 6 months | 345.0 | 441.9 | 0.78 (0.73-0.83) | <0.0001 |
| DLEU-related visits in 1 month | 35.5 | 46.2 | 0.77 (0.69-0.86) | <0.0001 |
| DLEU-related visits in 3 months | 103.2 | 126.8 | 0.81 (0.74-0.89) | <0.0001 |
| DLEU-related visits in 6 months | 168.5 | 202.0 | 0.83 (0.76-0.91) | <0.0001 |
| Debridement visits in 1 month | 21.5 | 26.5 | 0.81 (0.67-0.99) | 0.0357 |
| Debridement visits in 3 months | 60.1 | 76.9 | 0.78 (0.66-0.93) | 0.0060 |
| Debridement visits in 6 months | 94.1 | 121.6 | 0.77 (0.65-0.92) | 0.0036 |

### Economic Outcomes

The length of hospital stay days was 3% lower (statistically significant) in patients who had an application of STRAVIX compared to aCAMP patients. The index admission cost was 9% lower and the cost of all-cause visits was 25% lower within 1 month, 21% lower within 3 months and 20% lower within 6 months in STRAVIX patients compared to aCAMP patients. **(Table 4)**.

**Table 4:** Economic Outcomes.

| Outcome | STRAVIX | aCAMPs | Ratio (95% CI) | P-value |
| --- | --- | --- | --- | --- |
| Length of stay (days) | 12.3 | 12.8 | 0.97 (0.94-1.00) | 0.0326 |
| Index admission cost | \$42,263.61 | \$46,237.04 | 0.91 (0.89-0.94) | <0.0001 |
| Follow-up all-cause cost in 1 month | \$5,242.03 | \$6,977.46 | 0.75 (0.67-0.84) | <0.0001 |
| Follow-up all-cause cost in 3 months | \$12,936.57 | \$16,426.11 | 0.79 (0.73-0.85) | <0.0001 |
| Follow-up all-cause cost in 6 months | \$19,928.55 | \$24,803.49 | 0.80 (0.75-0.86) | <0.0001 |

## Discussion

Wound closure is often considered as the key parameter of clinical efficacy. However, associated outcomes, such as infection rate, rate of dehiscence, amputations, follow-up costs can all be pointers of successful management of surgical wounds. Use of CAMPs as an adjunct to standard surgical management of diabetic wounds has increased substantially over the last two decades. While a lot of evidence has been published on the efficacy of CAMPs in the outpatient setting, not enough data is available for the in-patient population. Head-to-head comparative data between different CAMPs is also lacking for management of surgical wounds. Real-world data analysis can therefore provide valuable insight into clinical outcomes after discharge, resource utilization and economic outcomes in management of DLEUs.

There is an ongoing debate whether regulatory pathway meaningfully predicts clinical effectiveness for CAMPs with respect to wound closure outcomes[17]. Results reported in this article span products of different regulatory pathways and demonstrate that 361 HCT/P products can be equivalent or better than 510(k) regulated products or PMA products. Due to lack of sufficient statistical difference in clinical outcomes between broad categories of CAMPs, decision makers should consider head-to-head comparative studies and real-world evidence between individual CAMPs to decide on which to use in practice.

The observation of decreased incidences of infection in this analysis is interesting as a recent article highlights the reduced incidences of infection in complex DFU patients who have had a STRAVIX application compared to reported rates for standard care[12]. Interestingly, in two different prospective studies with very similar patient population (complex diabetic surgical wounds, UT grade 2 and 3), reinfection rate in STRAVIX treated patients was much lower (7.5%) than what was reported in the study with an acellular CAMP (29.3%)[18]. The same aCAMP is also part of the aCAMP group in this study and hence the real-world data reported here aligns with previously published Level 1 studies.

A limitation of the study is the lack of a standard of care or a no-CAMPs group. The comparison to a “no skin substitute” control group was not logical as the patients who had an application of STRAVIX or other aCAMPs had more baseline comorbidities, and hence a completely different patient population. Higher percentage of patients who had a STRAVIX application had fat or muscle or bone exposed, had diabetes with complications, obesity, cellulitis, osteomyelitis and a higher Charlson comorbidity index. Patients who received STRAVIX could have also received negative pressure wound therapy (NPWT), which makes a direct comparison to no-CAMPs group difficult. This data also aligns with the current practice of selective use of CAMPs in hard-to-heal wounds which have a higher risk of complications. This finding is similar to another recent report where the authors reported that patients in the CAMP groups are generally more medically complex than standard of care group[9] and thus highlight a potential selection bias in which patients with more compromised physiology and at a higher risk of failure for wound closure, are more likely to receive CAMPs.

## Conclusion

Compared to the patients treated with some commonly available acellular CAMPs, the patients who received STRAVIX in addition to standard of care, had lower odds of DLEU related visit or debridement related visit, fewer adverse outcomes (infection reaction dehiscence, amputations after index admission discharge), a shorter length of hospital stay and lower total costs of the all-cause visits. Further randomized prospective studies with a head-to-head comparison can help validate these clinical and economic outcomes.

## Data Availability

All data produced in the present work are contained in the manuscript

## Acknowledgements

We would like to acknowledge Leo Nherera for his contribution in conceptualization and study design.

